# Association of Inflammation Biomarkers with Food Cravings and Appetite Changes Across the Menstrual Cycle

**DOI:** 10.1101/2023.01.30.23285198

**Authors:** Khushbu Agarwal, Alexis T. Franks, Xuemin Zhang, Enrique Schisterman, Sunni L. Mumford, Paule V. Joseph

**Affiliations:** Section of Sensory Science and Metabolism, National Institute on Alcohol Abuse and Alcoholism, National Institutes of Health, Department of Health and Human Services, Bethesda, Maryland, 20892, USA; National Institute of Nursing Research, National Institutes of Health, Department of Health and Human Services, Bethesda, Maryland, 20892, USA; Division of Statistical Analysis and Reporting (DSAR), Office of Research Reporting and Analysis (ORRA), Office of Extramural Research (OER), National Institutes of Health, Department of Health and Human Services, Besthesda, MD; Department of Biostatistics, Epidemiology and Informatics, University of Pennsylvania, Philadelphia, PA; Division of Population Health Research, Eunice Kennedy Shriver National Institute of Child Health and Human Development, Bethesda, MD

**Keywords:** Food cravings, Inflammation, Alcohol Intake, Stress, Body mass index, Menstrual Cycles

## Abstract

**Background:** Premenstrual symptoms, including food cravings, are often a regular complaint among menstruating women. However, existing evidence regarding the biological mechanisms by which these food cravings occur remains unclear. Inflammation may play an essential role in the occurence of these food cravings before menstruation.

**Purpose:** The purpose of the present study was to examine the associations between inflammatory markers and the risk of moderate/severe food cravings while accounting for changes in hormone levels and stress across the menstrual cycle.

**Methods:** The BioCycle Study followed women (n=259) aged 18-44 for two menstrual cycles. Food cravings (via questionnaire) were assessed up to four times per cycle. Each assessment corresponded to menses and mid-follicular, ovulation, and luteal phases of the menstrual cycle. A wide range of cytokine and chemokine levels (hsCRP, GCSF, GMCSF, IL-4, IL-6, RANTES, MIP1B, etc.) were assessed in blood samples collected at up to 8 visits per cycle, with visits timed using fertility monitors. Cravings for chocolate, sweets, salty, and other foods, and changes in appetite were determined to estimate the odds of moderate or severe cravings. Associations between inflammatory markers and risk of reporting a moderate/severe craving symptom at each cycle visit was determined using weighted generalized linear models (e.g., marginal structural models). Models were adjusted for age, BMI, and race, as well as time-varying covariates such as estradiol, stress, leptin, and total energy intake, and accounted for repeated measures (i.e., multiple cycles per woman). Both inflammatory markers and reports of cravings were modeled to account for variation at each visit.

**Results:** An association between higher inflammatory biomarkers such as hsCRP, GCSF, GMCSF, IL-4, IL-6, RANTES, MIP1B, and increased risk of moderate/severe cravings were identified across the menstrual cycle |all risk ratio>0.8, all CIs range>0.7-0.9|. hsCRP retained statistical significance after false discovery rate correction with chocolate, sweet, and salty cravings, while GCSF, GMCSF, IL-6, and RANTES retained significance with chocolate and sweet cravings only.

**Conclusion:** The results suggest a potential role of inflammation in food cravings and appetite changes across the menstrual cycle.

## 1. Introduction

Premenstrual symptoms are commonly experienced in women of reproductive age, and they significantly impact occupational performance and quality of life (Schoep, Adang, et al., 2019; Schoep, Nieboer, van der Zanden, Braat, & Nap, 2019). Previous studies show that approximately 90% of women report both physiological and psychological symptoms, including depressive mood, irritability, and food cravings, during the premenstrual phase of the menstrual cycle (luteal phase [days 16-28] of the menstrual cycle to the beginning of the next menses) (Chumpalova et al., 2020; Matsumoto, Asakura, & Hayashi, 2013). Most of these symptoms are thought to be caused by hormonal fluctuations and are exacerbated by psychological and behavioral factors (i.e., stress, depression, and eating disorders) (Fowler et al., 2019; Kroll-Desrosiers et al., 2017; Roney & Simmons, 2017). Food cravings have also been reported as a regular symptom among menstruating women (Hormes & Niemiec, 2017). Early studies showed an association between hormonal changes with food cravings during the menstrual cycle (Gorczyca et al., 2016; Hormes & Niemiec, 2017; Puder et al., 2006), though the mechanisms by which these changes occur remain poorly understood. Several studies have suggested a potential role of inflammation in premenstrual symptoms as inflammatory markers, like C-reactive protein (CRP) and several cytokines (tumor necrosis factor (TNF)-α and interleukin (IL)-6), fluctuate throughout the menstrual cycle (Bertone-Johnson et al., 2014; Blum et al., 2005; Capobianco et al., 2010; Lorenz, Worthman, & Vitzthum, 2015; Schliep et al., 2019; Wander, Brindle, & O’Connor, 2008; Whitcomb et al., 2014; Yama, Asari, Ono, Machida, & Miura, 2020). Associations between hsCRP and severity of premenstrual symptoms (Bertone-Johnson et al., 2014) and multiple risk factors for premenstrual syndrome (PMS) and premenstrual dysphoric disorder (PMDD), including depression, smoking, and high BMI (Puder et al., 2006; Yama et al., 2020) has been revealed amongst previous studies. Additionally, previous studies have demonstrated that anti-inflammatory agents relieve premenstrual symptoms (Gold, Wells, & Rasor, 2016; Lorenz et al., 2015).

However, the interplay of inflammation and food cravings during the menstrual cycle remains unknown. Studies need to consider the effects of fluctuating hormone levels on inflammatory markers and cravings. Thus, our present investigation was conducted using a cohort of healthy women from the BioCycle Study to examine the relationship of inflammation markers with food cravings and appetite changes while considering changing hormone and stress levels throughout the menstrual cycle.

## 2. Materials and Methods

### 2.1. Study Population

Participants included in the present study were women aged 18–44 who were regularly menstruating. Participants (n = 259) were recruited from the Western New York region and enrolled in the BioCycle Study (2005–2007), which followed participants for up to two complete menstrual cycles (Wactawski-Wende et al., 2009). Additional details regarding the recruitment process can be found in Wactawski-Wende et al. (2009). Participant eligibility criteria excluded women who used oral contraceptives during the preceding three months of the study, were pregnant, or were actively trying to conceive. Women with self-reported BMI between 18 and 35 kg/m^2^ at screening, a self-reported cycle length between 21 and 35 days for the past six months, and no history of gynecologic problems (i.e., endocrinologic disorders or known infertility) or chronic disease were included. The University at Buffalo Health Sciences Institutional Review Board approved the study and served as the institutional review board designated by the National Institutes of Health (NIH) for this study under a reliance agreement. All participants provided written informed consent.

### 2.2 Food Cravings and Change in Appetite (Outcome)

Food cravings were reported in a questionnaire regarding 17 menstrual symptoms and their severity in the previous week. The menstrual symptom questionnaire was completed at visits during menstruation and the mid-follicular, ovulation, and mid-luteal phases. Questions about food cravings were completed for 97% of visits, and included questions regarding cravings for chocolate, sweets, salty foods, other foods, and changes in the appetite (Gorczyca et al., 2016). Participants ranked their symptoms as none, mild, moderate or severe. We categorized severity as non/mild (reference group) or moderate/severe to estimate the odds of having a moderate or severe craving. We considered each food craving separately as well as any reported craving.

### 2.3 Inflammatory Markers (Exposure)

Eligible women who consented to participate provided fasting blood samples for a total of 16 visits (up to eight visits per cycle for up to two menstrual cycles). The first cycle visit began on the second day of menstruation. Visits were timed using Clearblue® Easy Fertility Monitors (Inverness Medical, Waltham, MA, USA) to correspond to each phase of the menstrual cycle (menstruation, midfollicular phase, late follicular phase, luteinizing hormone (LH) and follicle-stimulating hormone (FSH) surge, ovulation, and early-, mid-, and late-luteal phases). The fertility monitors specifically measured estrone-3-glucuronide and LH in urine to determine peak fertility. Other hormones, including total estradiol, progesterone, LH, FSH, and sex hormone binding globulin (SHBG) were measured in serum using Immulite 2000 Solid Phase competitive chemiluminescent enzymatic immunoassay (CLEIA; Siemens). hs-CRP was measured using the IMMULITE 2000 platform (Gaskins et al., 2012). Details regarding specimen collection and the timing of each visit were previously described Wactawski-Wende et al. (2009).

We measured cytokine and chemokine levels using BioSource 30-plex human cytokine assays (Invitrogen Corporation, Carlsbad, CA). A large scale of cytokines known to be involved in both menstrual cycle function and implantation (Vetrano, Wegman, Koes, Mehta, & King, 2020; Whitcomb et al., 2014) were covered in this standard panel. Especially: epithelial growth factor (EGF), eotaxin, fibroblast growth factor-basic (FGF-b), Granulocyte colony stimulating factor (G-CSF), Granulocyte-macrophage colony-stimulating factor (GM-CSF), hepatocyte growth factor (HGF), interferon (IFN) alpha (α), IFN-gamma(γ), IL-1β, IL-1 receptor antagonist (RA), IL-2, IL-2R, IL-4, IL-5, IL-6, IL-7, IL-8, IL-10, IL-12 (p40/p70), IL-13, IL-15, IL-17, interferon γ inducible protein (IP)-10, Monocyte Chemoattractant Protein(MCP)-1, monokine induced by interferon γ (MIG), Macrophage Inflammatory Protein (MIP)-1α, MIP-1β, Regulated upon Activation, Normal T Cell Expressed and Presumably Secreted (RANTES), TNF-α, and vascular endothelial growth factor (VEGF). All assys were run as per the manufacturor’s instructions. Concentrations were measured using the Luminex 100 IS system (Luminex Corp, Austin, TX). Detailed descrption on batches run can be found in our previous papers (Gaskins et al., 2012; Whitcomb et al., 2014).

### 2.4 Covariates

Age, race, smoking status, alcohol intake intensity, and reproductive history were obtained at baseline using standard questionnaires (Wactawski-Wende et al., 2009). Physical activity was assessed using the International Physical Activity Questionnaire(Craig et al., 2003). At the baseline visit, a trained research assistant measured height and weight (used to calculate BMI), and waist circumference at the natural waist using standardized protocols. Stress at baseline was assessed using the14 question Perceived Stress Scale (PSS-14), and during the cycle at the menstruation, mid-follicular, ovulation, and mid-luteal visits using the 4 question short version (Cohen, Kamarck, & Mermelstein, 1983; Schliep et al., 2015).

### 2.5 Statistical Analysis

All analyses were completed using SAS (version 9.4; SAS Institute, Cary, NC). Demographic characteristics were compared between those with any reported moderate or severe cravings or any moderate or severe changes in appetite during the study. We also evaluated the percentage of participants who reported moderate/severe cravings during each phase of the menstrual cycle. T-tests and chi-square tests were used for comparisons as appropriate.

Serum inflammation and cytokine levels were rightly skewed, and a natural log transformation was used to improve normality and meet model assumptions regarding the distribution of the residuals. We estimated associations between inflammatory markers and risk of reporting a moderate/severe craving symptom at each cycle visit using weighted generalized linear models (e.g., marginal structural models) that adjusted for age, BMI, race, as well as time-varying covariates of estradiol, stress, leptin, and total energy intake to account for changes in these factors across the menstrual cycle. Specifically, stabilized inverse probability weights were used to address the relations among hormone levels across the cycle; these models make use of longitudinal data to address the potential biases due to factors that are simultaneously intermediates and time-varying confounders (Cole & Hernán, 2008; Robins, Hernán, & Brumback, 2000). Results were adjusted for multiple comparisons using the false discovery rate (FDR). An alpha of <0.05 was considered statistically significant.

## 3. Results

### 3.1 Participant Characteristics and Changes in Cravings Across the Menstrual Cycle

Participants (n=259) included in the present study had a mean age of 27.3 years. The majority of the participants were of white race (59.3%) with a mean BMI of 24.1 kg/m^2^ (**Table 1**). About 57% of women reported any moderate/severe cravings and 36% reported moderate/severe changes in appetite during the study. Overall, most demographic characteristics were not associated with the report of moderate/severe cravings or changes in appetite, including age, race, BMI, physical activity, total energy intake, alcohol intensity, income, and parity. However, we did observe higher PSS scores among those who reported moderate/severe cravings or changes in appetite, and fewer women with less than a high school education reported changes in appetite. As expected, comparisons of symptom severity revealed that cravings (chocolate, sweet, salty) and appetite changes varied throughout the menstrual cycle, with the highest percentage of participants reporting moderate to severe symptoms during the premenstrual week (**Table 2**). Also, a greater percentage of women reported elevated craving severity for chocolate (24%) and sweet foods (22%) and salty foods (14%) during the premenstrual week (**Table 2**).

**Table 1:** Demographic characteristics of women in the BioCycle Study by severity of craving and appetite changes

|  | Total | Any cravings<br>(moderate/severe) |  |  | Any Change in Appetite<br>(moderate/severe) |  |  |
| --- | --- | --- | --- | --- | --- | --- | --- |
|  |  | Yes | No | p-value | Yes | No | p-value |
| N women | 259 | 148<br>(57) | 111<br>(43) |  | 94 (36) | 165<br>(64) |  |
| Age (years) <sup>2</sup> | 27.3 ±<br>8.2 | 27.4 ±<br>8.3 | 27.1 ±<br>8.1 | 0.7771 | 27.6 ±<br>8.7 | 27.1 ±<br>7.9 | 0.6332 |
| Race [*n* (%)] |  |  |  |  |  |  |  |
| White | 154<br>(59) | 90 (61) | 64 (58) | 0.8804 | 60 (64) | 94 (57) | 0.5831 |
| Black | 51<br>(20) | 28 (19) | 23 (21) |  | 16 (17) | 35 (21) |  |
| Other | 54<br>(21) | 30 (20) | 24 (22) |  | 18 (19) | 36 (22) |  |
| ≤High School Education [*n* (%)] | 33<br>(13) | 16 (11) | 17 (15) | 0.3469 | 4 (4) | 29 (18) | 0.0017 |
| Current Smoker [*n* (%)] | 10 (4) | 6 (4) | 4 (4) | 1 | 4 (4) | 6 (4) | 1 |
| Physical Activity [*n* (%)] |  |  |  |  |  |  |  |
| Low | 25 (9) | 13 (9) | 12 (11) | 0.331 | 8 (9) | 17 (10) | 0.1533 |
| Moderate | 92<br>(36) | 48 (32) | 44 (40) |  | 27 (29) | 65 (39) |  |
| High | 142<br>(55) | 87 (59) | 55 (50) |  | 59 (63) | 83 (50) |  |
| BMI, kg/m<sup>2</sup> | 24.1 ±<br>3.9 | 24.5 ±<br>3.9 | 23.6 ±<br>3.8 | 0.064 | 24.3 ±<br>3.3 | 24 ±<br>4.2 | 0.5228 |
| Waist to Hip Ratio | 0.75 ±<br>0.06 | 0.75 ±<br>0.06 | 0.75 ±<br>0.05 | 0.2175 | 0.76 ±<br>0.07 | 0.75 ±<br>0.05 | 0.3245 |
| Nulliparous [*n* (%)] | 187<br>(74) | 103<br>(72) | 84 (76) | 0.4724 | 68 (73) | 119<br>(74) | 0.8823 |
| Total Calories, kcal | 1607 ±<br>354 | 1629 ±<br>364 | 1579 ±<br>341 | 0.2624 | 1607 ±<br>381 | 1608 ±<br>340 | 0.9845 |
| Alcohol intensity |  |  |  |  |  |  |  |
| Never | 87<br>(34) | 42 (38) | 45 (30) | 0.351 | 30 (32) | 57 (35) | 0.5564 |
| Sometimes | 153<br>(59) | 63 (57) | 90 (61) |  | 55 (59) | 98 (59) |  |
| Often | 19 (7) | 6 (5) | 13 (9) |  | 9 (10) | 10 (6) |  |
| Income |  |  |  |  |  |  |  |
| Less than \$19,999 | 55<br>(21) | 36 (25) | 19 (17) | 0.4028 | 21 (23) | 34 (21) | 0.484 |
| \$20,000-\$39,999 | 61<br>(24) | 33 (23) | 28 (25) | | 19 (20) | 42 (26) | |
| \$40,000-\$74,999 | 72<br>(28) | 38 (26) | 34 (31) | | 23 (25) | 49 (30) | |
| \$75,000-\$99,999 | 45<br>(18) | 28 (19) | 17 (15) | | 21 (23) | 24 (15) | |
| \$100,000 or over | 24 (9) | 11 (8) | 13 (12) | | 9 (10) | 15 (9) | |
| Stress | 20.1 ±<br>6.8 | 21.4 ±<br>6.9 | 18.4 ±<br>6.1 | 0.0005 | 21.7 ±<br>7 | 19.3 ±<br>6.4 | 0.0063 |

**Table 2:** Prevalence of moderate/severe cravings symptoms across phases of the menstrual cycle

| N (%)* | **Appetite**<br>**Changed** | **Any**<br>**cravings** | **Chocolate**<br>**Cravings** | **Sweet**<br>**Cravings** | **Salty**<br>**Cravings** | **Other**<br>**Food**<br>**Cravings** |
| --- | --- | --- | --- | --- | --- | --- |
| **Premenstrual week** | 97 (19) | 171 (34) | 119 (24) | 112 (22) | 72 (14) | 49 (10) |
| **Menses** | 38 (8) | 99 (20) | 61 (12) | 64 (13) | 35 (7) | 21 (4) |
| **Follicular phase** | 24 (5) | 49 (10) | 23 (5) | 23 (5) | 22 (4) | 14 (3) |
| **Ovulatory phase** | 46 (10) | 72 (15) | 41 (9) | 34 (7) | 34 (7) | 26 (5) |

\*Number of cycles (%) with moderate/severe symptoms at each cycle phase

### 3.2 Inflammation Markers, and Craving Severity, and Appetite Changes

We observed an association between higher inflammatory biomarkers such as hsCRP, GCSF, GMCSF, IL-4, IL-6, RANTES, MIP1B, and increased risk of moderate/severe cravings across the menstrual cycle after adjusting for both time-fixed factors, as well as time-varying factors in weighted marginal structural models (**Table 3**). Specifically, increases in hsCRP, GCSF, GMCSF, HGF, IL-4, 6, and 13, RANTES, and MIG levels were associated with an increased risk of chocolate and sweet cravings. Salty food craving severity was only related to hsCRP, IFNA, and MIP levels. Similar to food craving severity, inflammation marker levels, including hsCRP, GM-CSF, HGF, IL-10 and 12, and RANTES were associated with changes in appetite (**Table 3**). Some of the associations retained statistical significance after false discovery rate correction; hsCRP, GCSF, GMCSF, IL-6, and RANTES with chocolate and sweet craving severity, and hsCRP with salty craving severity (**Supplemental Table S1**).

**Table 3:**
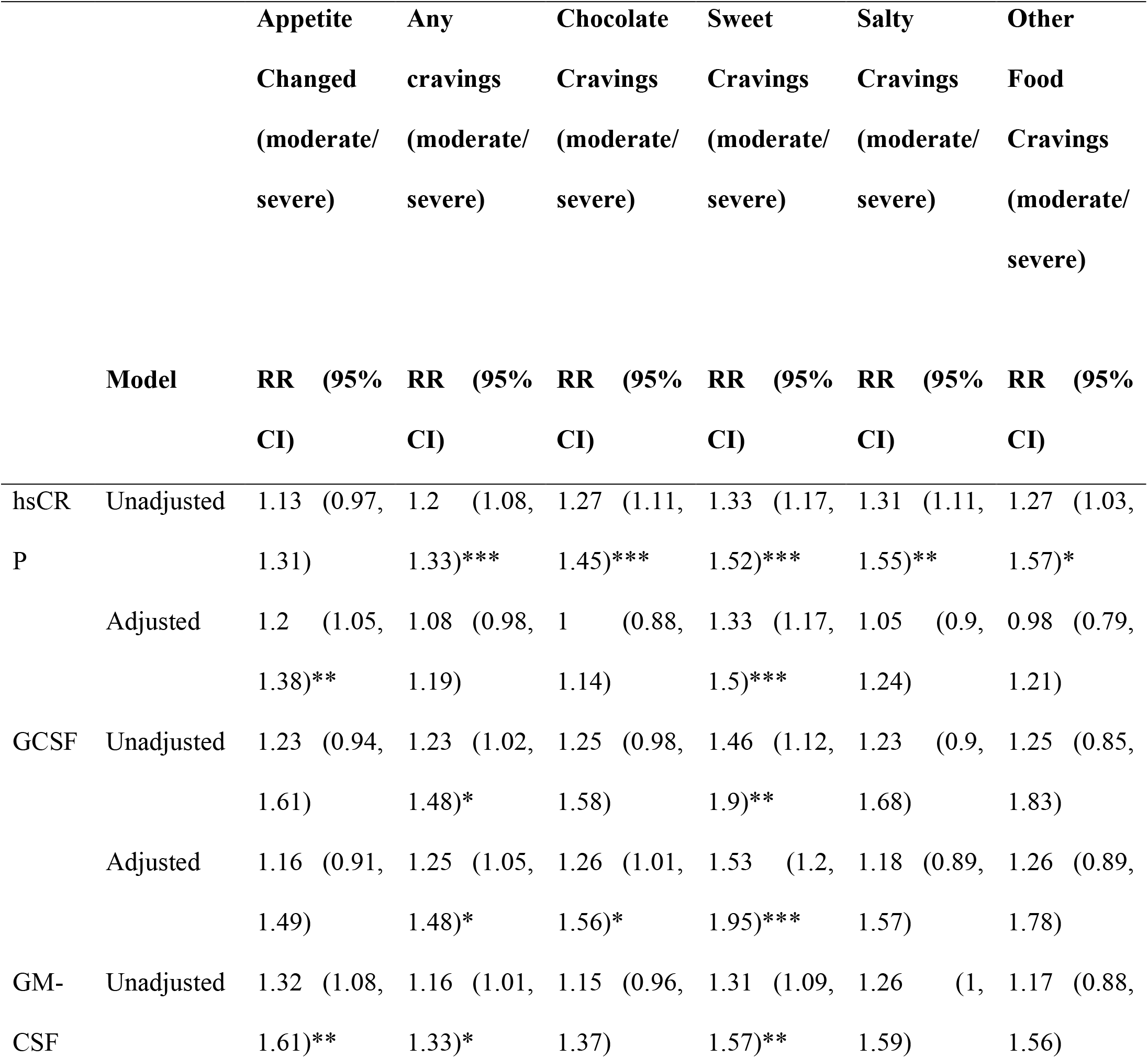
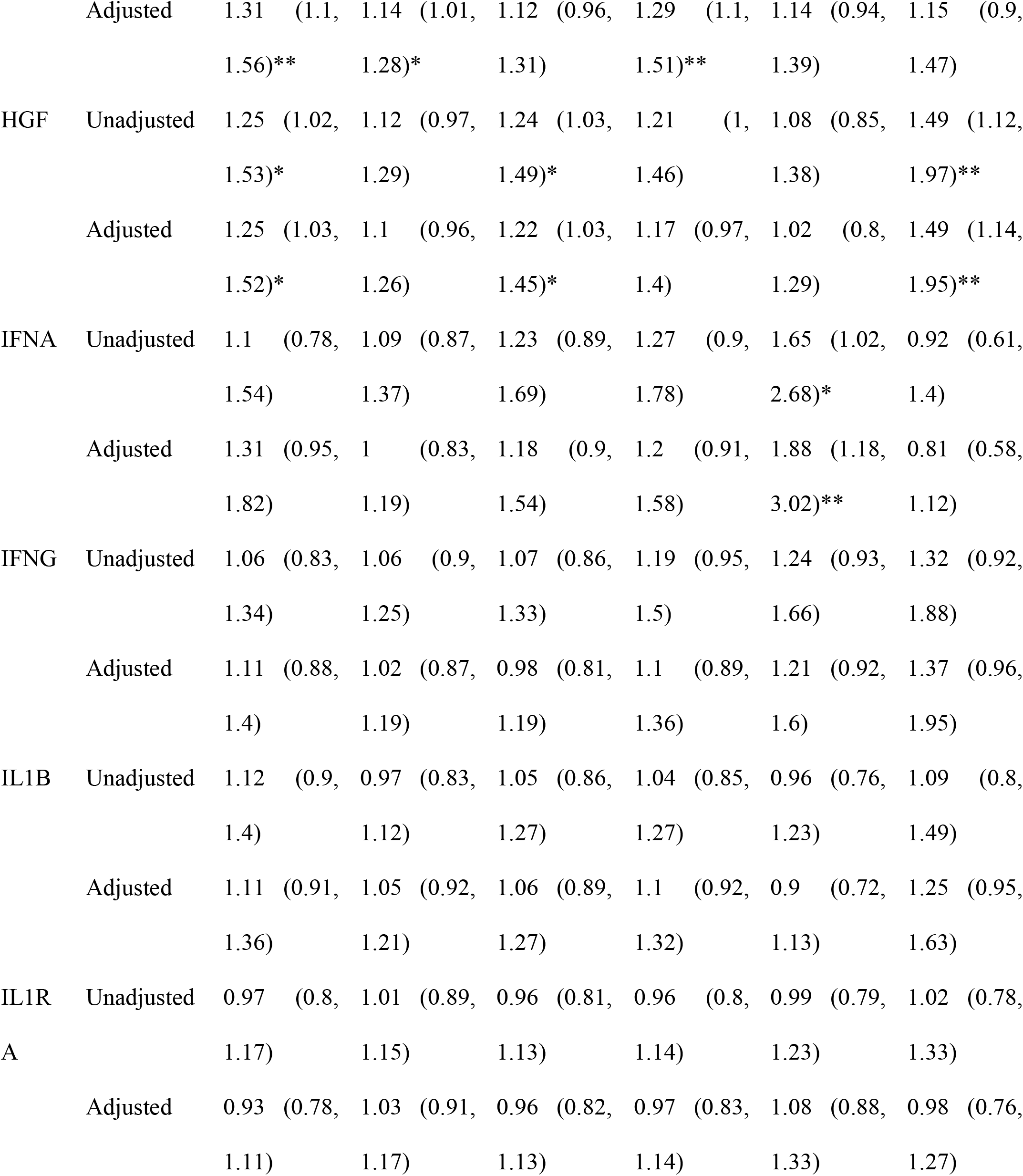
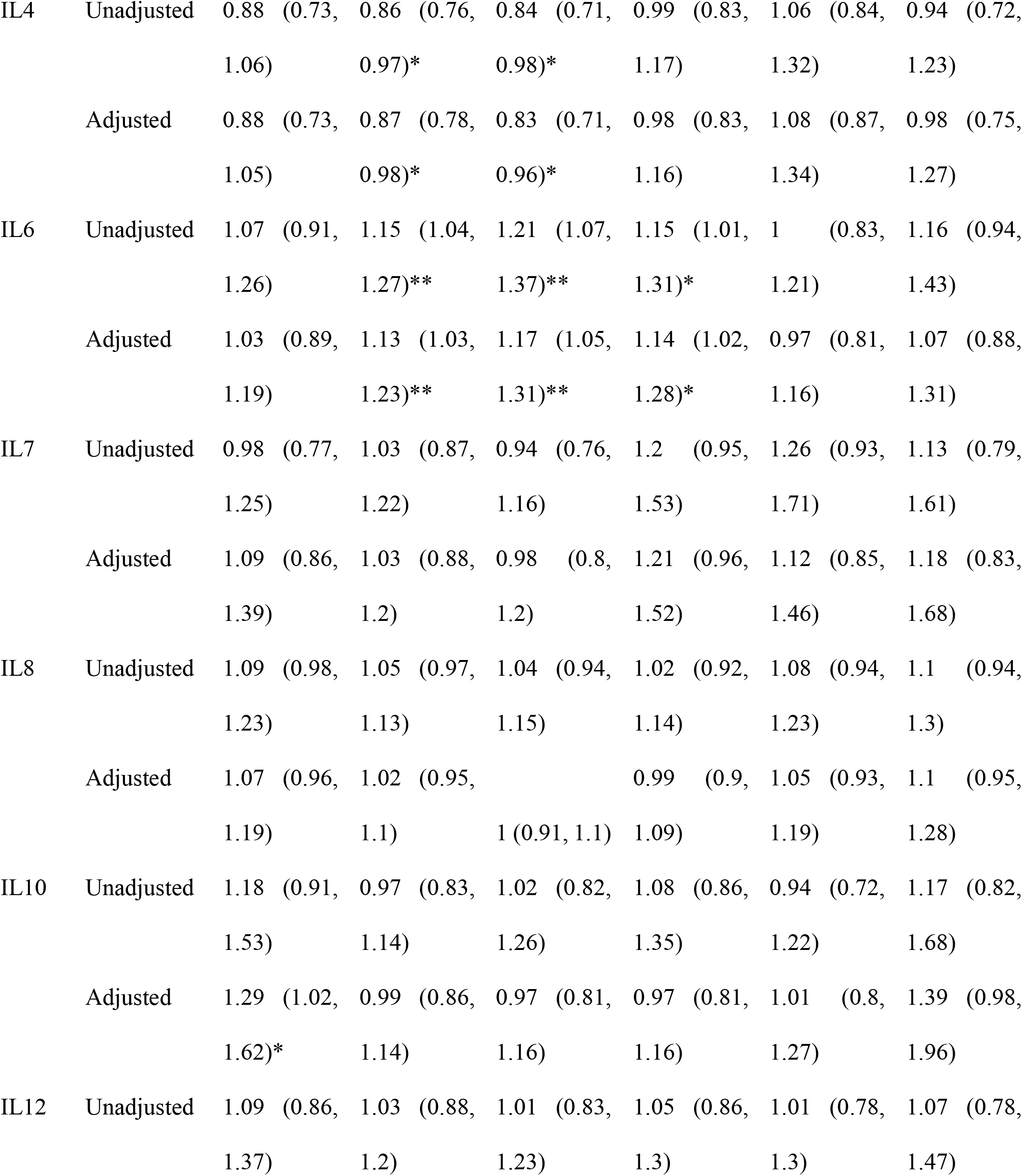
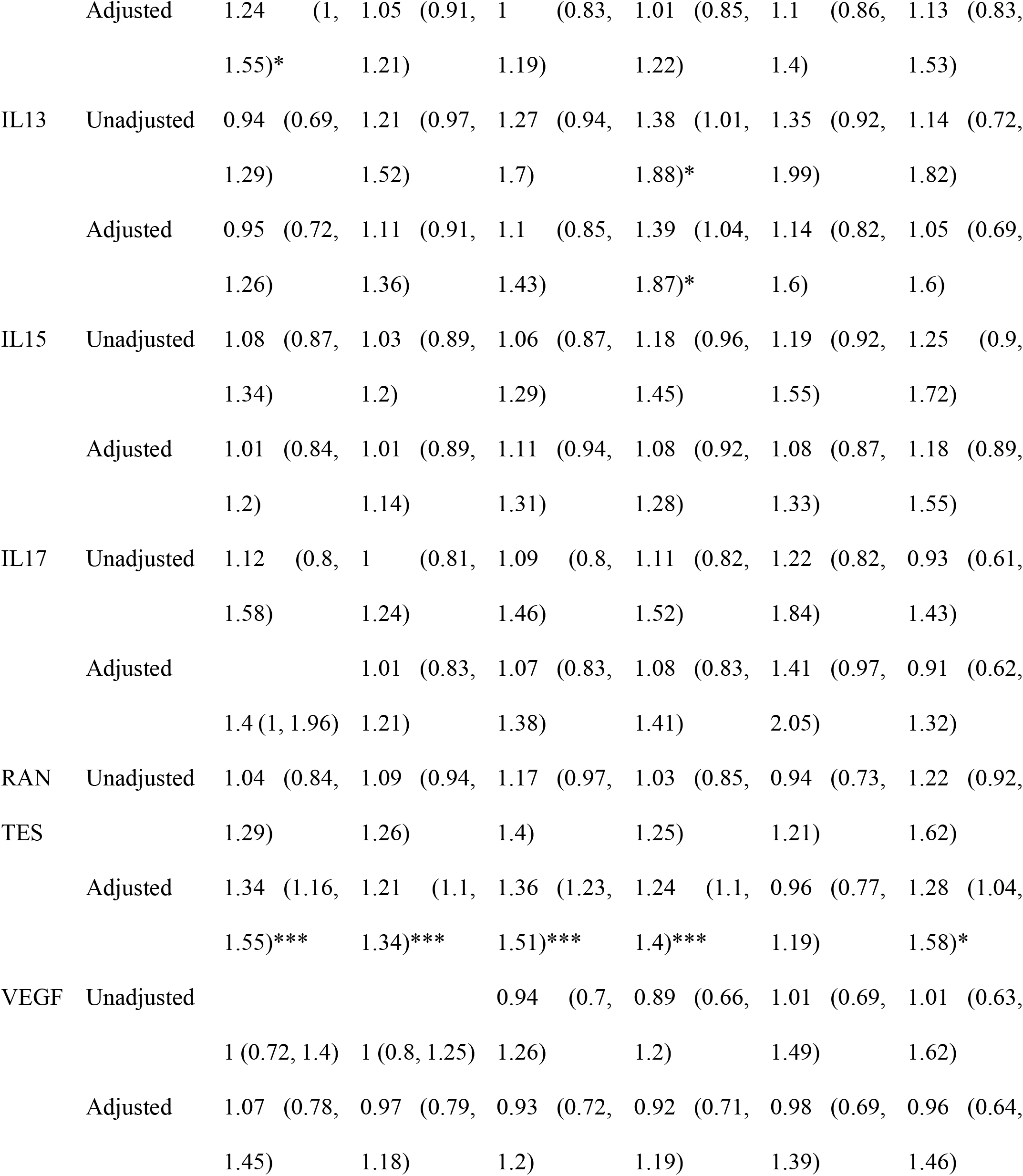
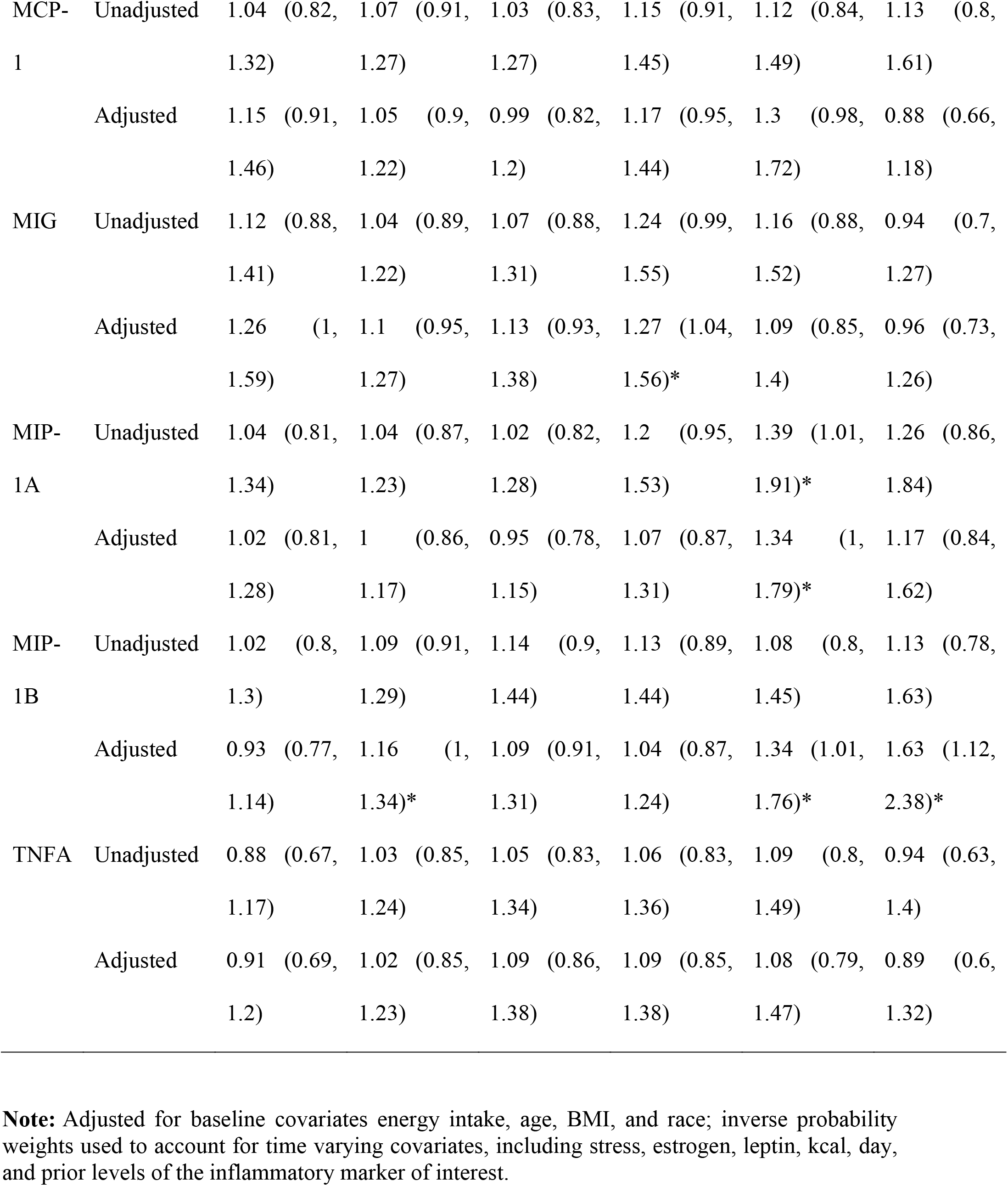
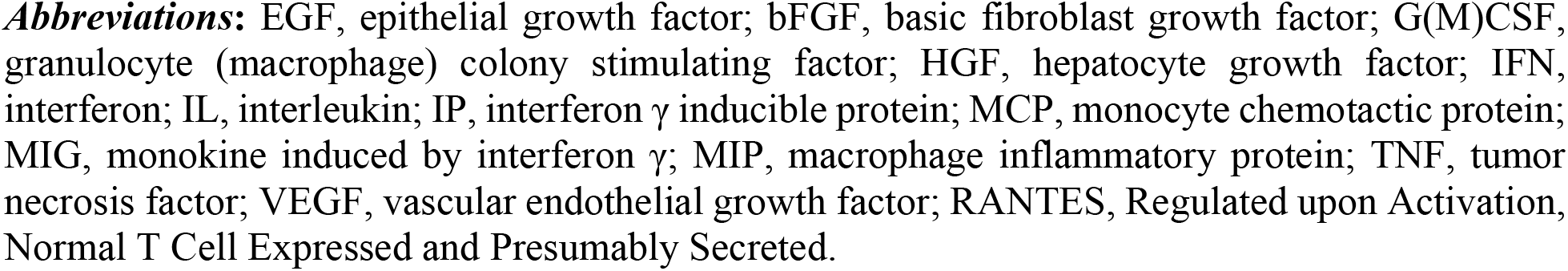
Associations between cytokines and inflammatory markers and presence of any moderate or severe cravings symptoms or changes in appetite.

## 4. Discussion

We found that inflammatory markers were associated with the severity of food cravings and appetite changes in women across the menstrual cycle, while considering fluctuating hormone levels. As expected and reported in our previous report (Gorczyca et al., 2016), cravings for chocolate and sweets were frequently reported throughout the menstrual cycle, with highest reports occurring during the premenstrual phase. Overall, these findings highlight the potential importance of inflammation in food cravings and appetite changes over the menstrual cycle.

The largely known biological explanation for the increased premenstrual food cravings, particularly carbohydrates, is related to hormonal fluctuations, i.e., increased progesterone levels during the luteal phase, mood fluctuations, and elevated stress levels. It has been demonstrated that sugar boosts the release of serotonin, a neurotransmitter that is known to modulate mood (Fernstrom & Wurtman, 1971; Teff, Young, & Blundell, 1989). The increased serotonin levels in the brain are known to be regulated by insulin (released as a consequence of carbohydrate intake), and the decline in insulin sensitivity may contribute to the occurrence of premenstrual symptoms (Escalante Pulido & Alpizar Salazar, 1999; Ezenwaka, Akanji, Adejuwon, Abbiyesuku, & Akinlade, 1993; Valdes & Elkind-Hirsch, 1991). In addition to these mechanisms, our results suggest that inflammation may also play an important role in premenstrual food and appetite changes.

A notable observation in our study was the association of elevated plasma levels of inflammatory markers (hsCRP, GCSF, GMCSF, IL-4, 6, 13 and RANTES) and craving severity. Inflammation has been extensively studied to be related to the severity of premenstrual symptoms (Granda, Szmidt, & Kaluza, 2021), with some reporting the strongest associations of plasma markers, hsCRP with mood and pain symptoms, appetite cravings/weight gain/bloating, abdominal cramps/back pain, and breast pain (Gold et al., 2016; Puder et al., 2006), and IL-2, IL-4, IL-10, IL-12, and IFN-γ with the severity of emotional and physical symptoms (Bertone-Johnson et al., 2014). Some have reported increased chemokine (RANTES) levels to be associated with menstrual cycle-associated symptoms (Roomruangwong, Sirivichayakul, Carvalho, & Maes, 2020). Our results corroborate previous literature evidence suggesting inflammation markers are associated with craving severity as a premenstrual symptom in women.

To date, there is scant evidence that depicts the existing relationship between inflammation, food cravings and appetite changes during menstruation, despite the usage of anti-inflammatory medications by some women to relieve their symptoms. Since studies conducted across the last few decades provide clear evidence of the role of dopamine in increasing food cravings and the elevation in dopamine levels caused by systemic inflammation, understanding the relationship of a wide range of inflammation markers with food cravings and appetite changes would enable future indepth studies on understanding the moderating effect of inflammation on elevated dopamine and food cravings during menstruation cycle phases.

## 5. Strengths and Limitations

Some of the strengths of the present study are (i) the use of multiple inflammatory markers, including hsCRP as a global marker of systemic inflammation and a large panel of cytokines, to assess the relationship of inflammation with elevated food cravings and appetite changes during the menstrual cycle phases, (ii) large sample and repeated measure collection in terms of days and menstrual cycles. There are also some limitations of the present study; since the study design is observational, this may not allow us to establish a causal relationship between increased plasma inflammatory marker levels and craving severity as a pre-menstruation symptom. However, we are accounting for changes in hormones and stress over the cycle that influence both cravings and inflammation using appropriate causal inference methods, and we still see an association here. Also the percentage of women reporting high levels of stress was relatively less than reports based on the national survey (“ American Psychological Association. Stress in America Findings 2022: Stress by Gender,” [Accessed 10 December 2022]). However, it is essential to note that in the current investigation, the women self-reported normal menstrual cycles, which could actually be related to the lower rates of stress as those with high stress may have impaired ovulatory function and irregular periods. So overall, they are healthy and regularly menstruating, which may limit generalizability. Also, these women were not taking oral contraceptives. One of the other limitations of the present study is the lack of direct measures for food cravings which need to be considered in future studies for replication purposes.

## 6. Conclusion

Overall, we observed associations between various inflammatory markers with an increased risk of moderate/severe cravings and changes in appetite during the menstrual cycle. These findings importantly account for fluctuating hormones and stress levels over the cycle. This points to the important role of inflammation in food cravings and appetite changes across the menstrual cycle phases. The present study results highlight the importance of future in-depth mechanistic studies to understand the moderating effect of inflammation on the link between elevated dopamine and food cravings during menstruation as reported across the literature.

## Data Availability

All data produced in the present study are available upon reasonable request to the authors

