## Supplemental TableS1 for "Association of Inflammation Biomarkers with Food Cravings and Appetite Changes Across the Menstrual Cycle"

**SUPPLEMENTAL TABLE S1:** Results of False Discovery Rate (FDR) correction for associations between cytokines and inflammatory markers and presence of any moderate or severe cravings symptoms or changes in appetite.

|  |  | **Appetite Changed (moderate/severe)** | **Any cravings (moderate/severe)** | **Chocolate Cravings (moderate/severe)** | **Sweet Cravings (moderate/severe)** | **Salty Cravings (moderate/severe)** | **Other Food Cravings (moderate/severe)** |
| --- | --- | --- | --- | --- | --- | --- | --- |
|  | **Model** | **p-value** | **p-value** | **p-value** | **p-value** | **p-value** | **p-value** |
| hsCRP | Unadjusted | 0.6503 | 0.011* | 0.0123* | 0.0005* | 0.0397* | 0.3064 |
|  | Adjusted | 0.0641 | 0.4744 | 0.9841 | 0.0003* | 0.7028 | 0.9022 |
| GCSF | Unadjusted | 0.6503 | 0.1722 | 0.3588 | 0.037* | 0.4992 | 0.6573 |
|  | Adjusted | 0.4616 | 0.0883 | 0.1915 | 0.0048* | 0.7028 | 0.546 |
| GM-CSF | Unadjusted | 0.1672 | 0.1983 | 0.4109 | 0.037* | 0.3072 | 0.6573 |
|  | Adjusted | 0.0258 | 0.1724 | 0.5787 | 0.0104* | 0.6321 | 0.5505 |
| HGF | Unadjusted | 0.3569 | 0.4514 | 0.1571 | 0.2197 | 0.8027 | 0.1365 |
|  | Adjusted | 0.1453 | 0.5257 | 0.1343 | 0.266 | 0.9416 | 0.0779 |
| IFNA | Unadjusted | 0.8508 | 0.8721 | 0.5748 | 0.3455 | 0.3072 | 0.8278 |
|  | Adjusted | 0.2728 | 0.9862 | 0.5912 | 0.4416 | 0.2013 | 0.546 |
| IFNG | Unadjusted | 0.8508 | 0.8721 | 0.8159 | 0.2817 | 0.4913 | 0.6573 |
|  | Adjusted | 0.6253 | 0.9862 | 0.9841 | 0.6345 | 0.6321 | 0.3815 |
| IL1B | Unadjusted | 0.8508 | 0.8721 | 0.8159 | 0.7806 | 0.9082 | 0.8278 |
|  | Adjusted | 0.5217 | 0.9726 | 0.902 | 0.5812 | 0.7028 | 0.4324 |
| IL1RA | Unadjusted | 0.8508 | 0.9832 | 0.8159 | 0.7478 | 0.9775 | 0.9327 |
|  | Adjusted | 0.6361 | 0.9726 | 0.9197 | 0.8544 | 0.7028 | 0.9022 |
| IL4 | Unadjusted | 0.6928 | 0.1423 | 0.1571 | 0.8763 | 0.8027 | 0.8278 |
|  | Adjusted | 0.3435 | 0.1603 | 0.1123 | 0.8742 | 0.7028 | 0.9022 |
| IL6 | Unadjusted | 0.8508 | 0.0705 | 0.03* | 0.1971 | 0.9775 | 0.6573 |
|  | Adjusted | 0.8103 | 0.0883 | 0.0747 | 0.0922 | 0.861 | 0.7466 |
| IL7 | Unadjusted | 0.9306 | 0.8721 | 0.8159 | 0.2817 | 0.4913 | 0.8278 |
|  | Adjusted | 0.6361 | 0.9862 | 0.9841 | 0.266 | 0.7028 | 0.6567 |
| IL8 | Unadjusted | 0.6503 | 0.6977 | 0.8159 | 0.774 | 0.6248 | 0.6573 |
|  | Adjusted | 0.4616 | 0.9726 | 0.9841 | 0.8742 | 0.7028 | 0.546 |
| IL10 | Unadjusted | 0.6951 | 0.8721 | 0.9064 | 0.7069 | 0.8027 | 0.8278 |
|  | Adjusted | 0.1453 | 0.9862 | 0.9841 | 0.8544 | 0.9533 | 0.3657 |
| IL12 | Unadjusted | 0.8508 | 0.8721 | 0.932 | 0.7478 | 0.9775 | 0.8278 |
|  | Adjusted | 0.154 | 0.9726 | 0.9841 | 0.8742 | 0.7028 | 0.6926 |
| IL13 | Unadjusted | 0.8508 | 0.3707 | 0.4027 | 0.1971 | 0.4913 | 0.8278 |
|  | Adjusted | 0.8103 | 0.7418 | 0.902 | 0.0933 | 0.7028 | 0.9022 |
| IL15 | Unadjusted | 0.8508 | 0.8721 | 0.8159 | 0.2817 | 0.4992 | 0.6573 |
|  | Adjusted | 0.9391 | 0.9862 | 0.5912 | 0.6345 | 0.7028 | 0.5505 |
| IL17 | Unadjusted | 0.8508 | 0.9916 | 0.8159 | 0.7069 | 0.6577 | 0.8278 |
|  | Adjusted | 0.154 | 0.9862 | 0.902 | 0.7243 | 0.3519 | 0.8194 |
| RANTES | Unadjusted | 0.8508 | 0.6977 | 0.3869 | 0.797 | 0.8027 | 0.6573 |
|  | Adjusted | 0.0023* | 0.0028* | <.0001* | 0.0048* | 0.8141 | 0.1566 |
| VEGF | Unadjusted | 0.9866 | 0.9916 | 0.8159 | 0.7069 | 0.9775 | 0.9596 |
|  | Adjusted | 0.8103 | 0.9862 | 0.902 | 0.7243 | 0.9416 | 0.9022 |
| MCP-1 | Unadjusted | 0.8508 | 0.8721 | 0.906 | 0.4259 | 0.7866 | 0.8278 |
|  | Adjusted | 0.4616 | 0.9726 | 0.9841 | 0.3284 | 0.3519 | 0.658 |
| MIG | Unadjusted | 0.8508 | 0.8721 | 0.8159 | 0.2206 | 0.6248 | 0.8278 |
|  | Adjusted | 0.154 | 0.5334 | 0.5912 | 0.0922 | 0.7028 | 0.9022 |
| MIP-1A | Unadjusted | 0.8508 | 0.8721 | 0.906 | 0.2817 | 0.3072 | 0.6573 |
|  | Adjusted | 0.9051 | 0.9862 | 0.902 | 0.7243 | 0.3519 | 0.6567 |
| MIP-1B | Unadjusted | 0.9306 | 0.8445 | 0.6495 | 0.5346 | 0.8027 | 0.8278 |
|  | Adjusted | 0.6361 | 0.186 | 0.7837 | 0.8544 | 0.3519 | 0.1389 |
| TNFA | Unadjusted | 0.8508 | 0.8721 | 0.8159 | 0.7478 | 0.8027 | 0.8278 |
|  | Adjusted | 0.6361 | 0.9862 | 0.902 | 0.7243 | 0.8141 | 0.7802 |
